# Sociodemographic inequalities related to the early beginning of antenatal care visits in Peruvian women: an analysis of the Demographic and Family Health Survey between 2019 and 2022

**DOI:** 10.1101/2024.05.12.24307249

**Authors:** Claudio Intimayta-Escalante

## Abstract

**Introduction:** Starting antenatal care within the first three months of pregnancy is crucial for maternal and fetal health, but sociodemographic barriers hinder timely care initiation for women. This study aims to assess sociodemographic inequalities in the initiation of antenatal care visits among Peruvian women.

**Methods:** A cross-sectional analysis with data from the 2019-2022 Demographic and Family Health Survey in Peru was conducted. Weighted Cox regression models helped calculate adjusted Hazard Ratios (aHR), and the Slope Index of Inequality (SII) was used to measure how sociodemographic factors like age, education, location, insurance, and ethnicity influenced the timing of antenatal care initiation.

**Results:** The study included 22668 Peruvian women aged 18 to 49. Among these women, the mean age was 31.45 years. Only 30.63% of women started their antenatal care visits in the first month of pregnancy. Additionally, women without education (aHR: 0.74, 95%CI: 0.63 to 0.85, p¡0.001), those in urban areas (aHR: 0.94, 95%CI: 0.89 to 0.98, p=0.003), and individuals of Quechua or Aymara descent (aHR: 0.91, 95%CI: 0.87 to 0.95, p¡0.001) were less likely to initiate antenatal care in the first months. Furthermore, individuals aged 18 to 29 (SII: -0.22, 95%CI: -0.26 to -0.18, p¡0.001), those without education (SII: -0.03, 95%CI: -0.04 to -0.02, p¡0.001), residing in rural areas (SII: -0.75, 95%CI: -0.78 to -0.71, p¡0.001), or living outside the capital (SII: -0.65, 95%CI: -0.70 to -0.60, p¡0.001) exhibited similar patterns.

**Conclussion:** Sociodemographic inequalities exist in the early beginning of antenatal care visits are evident among Peruvian women, especially impacting individuals in rural or non-capital regions with lower education levels and belonging to the Quechua or Aymara ethnic communities.

## INTRODUCTION

The World Health Organization (WHO) recommends the beginning of antenatal care visits within the first three months of pregnancy to safeguard maternal and fetal health (1). Despite Peru’s commendable achievement of over 80% coverage of antenatal care visits, several barriers persist, impeding timely initiation of care among women (2). Factors such as limited access to healthcare services, sociodemographic characteristics including age, health insurance status, and place of residence, alongside personal factors such as disinterest and experiences of partner violence, contribute to delayed initiation of antenatal care (3,4).

The urgency of addressing these barriers is underscored by the stark rise in maternal mortality rates in Peru (5). This because only in 2019, the nation recorded 302 maternal deaths, a figure that witnessed a concerning surge amidst the COVID-19 pandemic, soaring to 493 maternal deaths in 2021 (6). This escalating toll highlights the critical need for policies that prioritize early antenatal care, especially for economically disadvantaged women, as the absence of such measures exacerbates existing disparities rooted in socioeconomic conditions (7-9).

While there exists evidence highlighting disparities in sexual and reproductive health programs across South America, scant attention has been devoted to understanding variations in the initiation of antenatal care visits, particularly within the low and middle incomes countries (10-12). However, the gaps mediated for sociodemographic inequalities in the timing of antenatal care initiation among Peruvian women during gestation that increases in the COVID-19 pandemic (13). This study aims to assess sociodemographic inequalities in the early beginning of antenatal care visits.

## METHODS

### Study Design

An analytical cross-sectional study was developed, with data from the Demographic and Family Health Survey (DHS) between 2019 and 2022. The DHS is an annual survey developed throughout Peru, a Latin American country with approximately 32 million inhabitants (mainly concentrated in Lima, the capital of this country) (14). The study focused on evaluating the sociodemographic and health characteristics of Peruvian women aged 18 to 49 who had experienced a previous pregnancy and provided information on antenatal care visits.

### Evaluation of Antenatal Care Visits

The definition used for the early beginning of antenatal care visits was the time of the first antenatal care visit between nine months of gestation. Also, was assessed if a complete evaluation was conducted during antenatal care visits, including blood and urine tests, HIV/AIDS screening, blood pressure measurements, tetanus vaccination, iron supplementation, education on nutrition, and guidance on managing pregnancy complications (3,15).

### Sociodemographic Conditions

Additionally, was assessed sociodemographic factors that could influence the timing of antenatal care initiation during pregnancy among Peruvian women. Thus, was addressed characteristics such as age group (18 to 24, 25 to 34, 35 to 49 years old), educational level (without education, elementary, high school, or university), wealth index (first, second, third, fourth, and last quintile), area of residence (rural and urban), living in the capital (yes or no), ethnic group (white or mestizo, Quechua or Aymara, and Afro-Peruvian), and affiliation to health insurance during pregnancy (yes or no).

### Statistical Analysis

The statistical analysis was developed in R Studio v.4.2.2 (https://cran.r-project.org/), including the DHS complex sample design. Categorical variables were described by presenting their frequencies and percentages. It was calculated the average values and their corresponding 95% confidence intervals to describe the numerical variables. In the bivariate analysis, the Wald test was used to find the health and sociodemographic factors that make a difference in the average month of the first prenatal care visit between the groups that were looked at.

However, to address differences in the beginning of antenatal care visits, a survival analysis approach was used with the “coxphw” package (https://cran.r-project.org/web/packages/coxphw/coxphw.pdf), defining the follow-up time as the nine months of pregnancy and the event of interest as the month during gestation when antenatal care visits began. In this way, comparisons of the survival function were performed using Kaplan-Meier plots and the long-rank test with the “survfit” and “ggsurvplot” function, evaluating the cumulative percentage of pregnant women with antenatal care visits during the nine months of pregnancy. Also, was looked at the impact of the characteristics that were measured at the begining of antenatal care visits using weighted Cox regression models for complex samples. These models estimated the crude Hazard Ratio (cHR) and adjusted for the other variables (aHR).

### Inequality Analysis

The Slope Inequality Index (SII) was used to evaluate inequalities at the beginning of antenatal care visits (first, second, third, fourth, and fifth to ninth months of pregnancy). Thus, comparisons were made according to the categories of the different sociodemographic and health characteristics assessed. An SII value between -1 and 0 suggests greater inequality associated with the assessed socioeconomic condition, while a value between 0 and 1 indicates less inequality related to the socioeconomic characteristic under evaluation (16).

### Ethical Aspects

The study was developed by analyzing data from the DHS platform in Peru (https://proyectos.inei.gob.pe/microdatos/). This survey is developed with the informed consent of the participants. In addition, the study did not have information that would allow identification of the participants included in the research. Moreover, all ethical standards were strictly followed throughout the study.

## RESULTS

The study included 22668 Peruvian women aged 18 to 49 who usually lived where they were surveyed and had a previous pregnancy (Appendix 1). The mean age was 31.45 years (95% CI: 31.28 to 31.62). In addition, the majority had high school and university education (84.72%, 95%CI: 84.00 to 85.41), while almost a third were among the two highest wealth quintiles (34.88%, 95%CI: 33.64 to 36.15). Regarding residence, eight out of ten women evaluated lived in an urban area and six out of ten in regions outside of the capital (Table 1). Regarding ethnic identification, 48.39% identified as mestizo (95%CI: 47.30 to 49.48), followed by 24.34% as Quechua or Aymara (95%CI: 23.45 to 25.24), 10.51% as Afro-Peruvian (95%CI: 9.94 to 11.12), and 7.12% as White or Mestizo (95%CI: 6.58 to 7.70). In addition, six out of ten women reported having been affiliated to national health insurance during their pregnancy. In addition, almost all confirmed that a physician or obstetrician attended in their antenatal care visits, and only half received a complete evaluation in antenatal care (Table 1).

**Table 1.** Sociodemographic and health characteristics of Peruvian women with a history of pregnancy registered in the Demographic and Family Health Survey between 2019 and 2022.

| Characteristics | Peruvian women with ACVs |  | Month of start of ACVs |  |
| --- | --- | --- | --- | --- |
|  | n (%*) | 95%CI | Mean (95%Ci) | P-value** |
| **Age Group?** |  |  |  |  |
| 18 to 24 years old | 4309 (20.71) | 19.88 a 21.57 | 2.79 (2.72 a 2.86) | <0.001 |
| 25 to 34 years old | 11494 (43.43) | 42.42 a 44.45 | 2.32 (2.28 a 2.36) |  |
| 35 to 49 years old | 6865 (35.86) | 34.78 a 36.96 | 2.28 (2.23 a 2.32) |  |
| **Educational Level?** |  |  |  |  |
| Without education | 295 (0.99) | 0.84 a 1.17 | 2.85 (2.62 a 3.07) | <0.001 |
| Elementary | 3987 (14.28) | 13.62 a 14.97 | 2.74 (2.67 a 2.81) |  |
| High School | 10943 (47.1) | 46.04 a 48.16 | 2.54 (2.5 a 2.58) |  |
| University | 7443 (37.62) | 36.51 a 38.75 | 2.08 (2.04 a 2.13) |  |
| **Wealth Index?** |  |  |  |  |
| First Quintile (Q1) | 1937 (15.85) | 14.88 a 16.86 | 1.9 (1.83 a 1.98) | <0.001 |
| Q2 | 2977 (19.04) | 18.12 a 19.99 | 2.23 (2.15 a 2.31) |  |
| Q3 | 4491 (22.09) | 21.21 a 23 | 2.36 (2.3 a 2.42) |  |
| Q4 | 6683 (24.93) | 23.98 a 25.9 | 2.6 (2.55 a 2.66) |  |
| Last Quintile (Q5) | 6580 (18.1) | 17.37 a 18.85 | 2.78 (2.73 a 2.83) |  |
| **Residence Area?** |  |  |  |  |
| Rural | 6952 (17.7) | 16.93 a 18.51 | 2.68 (2.63 a 2.73) | 0.191 |
| Urban | 15716 (82.3) | 81.49 a 83.07 | 2.34 (2.31 a 2.37) |  |
| **Do you live in the Capital?** |  |  |  |  |
| No | 19544 (61.3) | 59.95 a 62.63 | 2.51 (2.49 a 2.54) | 0.565 |
| Yes | 3124 (38.7) | 37.37 a 40.05 | 2.22 (2.17 a 2.28) |  |
| **Ethnic Group?** |  |  |  |  |
| White or Mestizo | 10686 (58.79) | 57.72 a 59.84 | 2.26 (2.22 a 2.3) | 0.020 |
| Quechua or Aymara | 7716 (27.58) | 26.65 a 28.54 | 2.62 (2.56 a 2.67) |  |
| Afro-Peruvians | 2300 (11.14) | 10.52 a 11.78 | 2.53 (2.46 a 2.61) |  |
| Others | 767 (2.49) | 2.2 a 2.83 | 2.49 (2.29 a 2.69) |  |
| **Were you affiliated to SIS during pregnancy?** |  |  |  |  |
| No | 4530 (31.74) | 30.54 a 32.96 | 2.06 (2 a 2.12) | <0.001 |
| Yes | 14230 (68.26) | 67.04 a 69.46 | 2.53 (2.5 a 2.57) |  |
| **Was the ACVs performed by a physician or obstetrician?** |  |  |  |  |
| No | 979 (3.05) | 2.74 a 3.4 | 2.78 (2.64 a 2.92) | 0.317 |
| Yes | 21689 (96.95) | 96.6 a 97.26 | 2.39 (2.36 a 2.42) |  |
| **Did you have ACVs with complete evaluations?** |  |  |  |  |
| No | 8100 (49.57) | 48.33 a 50.8 | 2.47 (2.42 a 2.52) | 0.666 |
| Yes | 8153 (50.43) | 49.2 a 51.67 | 2.26 (2.22 a 2.31) |  |

**95%CI:** 95% Confidence Interval, **ACVs:** Antenatal Care Visits.
\*Weighted percentage for complex sample.
\*\*P-value estimated by Wald test.

Only 30.63% (95%CI: 29.66 to 31.63) had their antenatal care visit in the first month of pregnancy (Figure 1). While half started their antenatal care visits between the second and fourth months of pregnancy (60.75%), Thus, it was identified that characteristics such as age group, educational level, wealth index, ethnicity, and health insurance during pregnancy mediated differences in the average month when they started antenatal care visits (Table 1).

**Figure 1.**
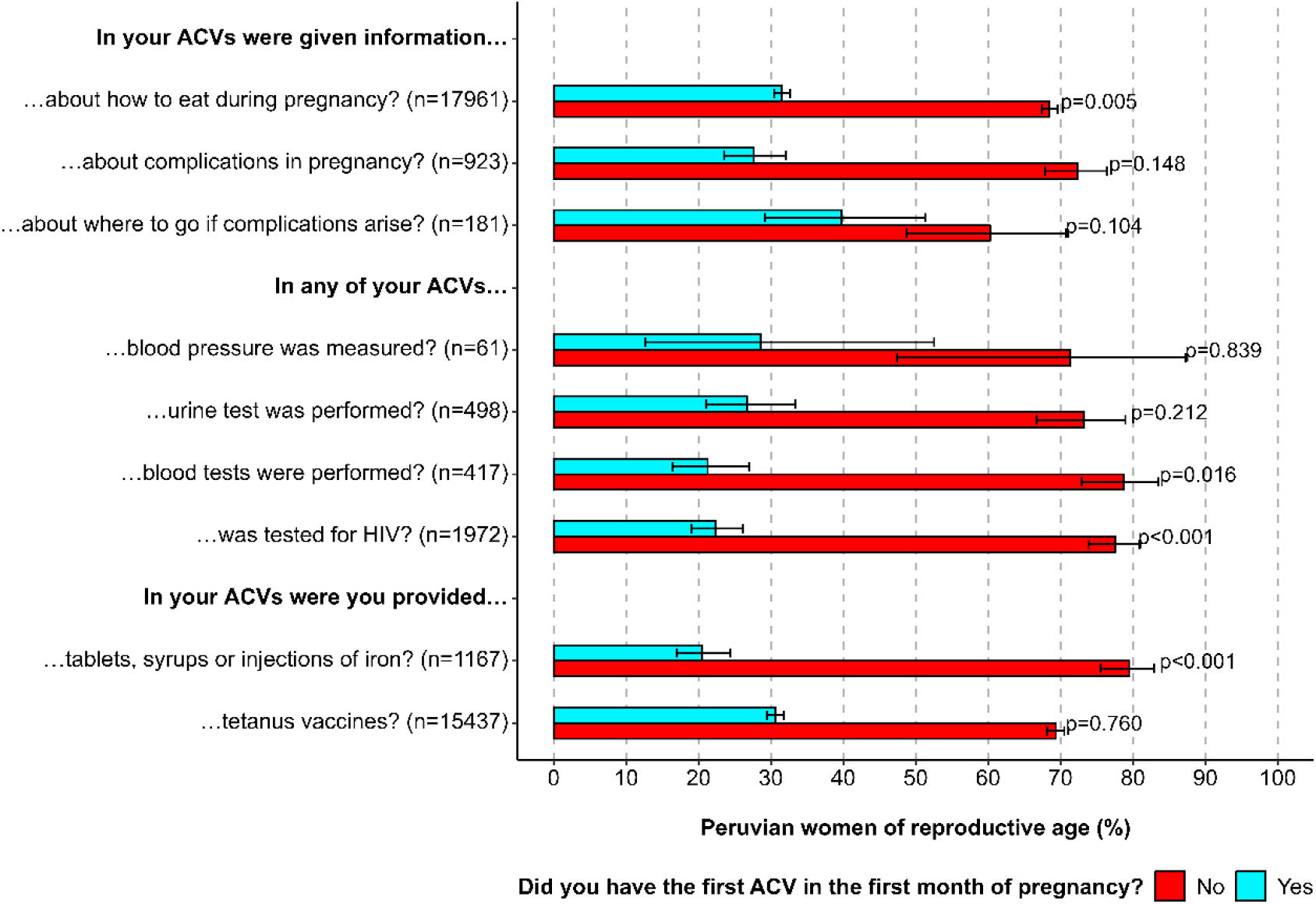
Evaluations to be completed at antenatal care visits in Peruvian women **ACVs:** Antenatal Care Visits.

Among the Peruvian women who received a complete evaluation in their antenatal care visits, 27.65% were told about pregnancy complications, and 39.73% were told where to go in case of any complications. In addition, it was found that 31.52% explained how to eat during pregnancy. Likewise, 28.64%, 26.75%, 26.75%, 21.28%, and 22.40% were evaluated for blood pressure, urine, blood, and HIV/AIDS. Also, 20.48% and 30.62% were provided with iron supplements and tetanus vaccine, respectively (Figure 1). The assessment revealed that sociodemographic and health factors significantly influenced when women began their antenatal care visits. Older women were more likely to begin antenatal care visits in the first months of pregnancy compared to those aged 18 to 24 years (Figure 2).

**Figure 2.**
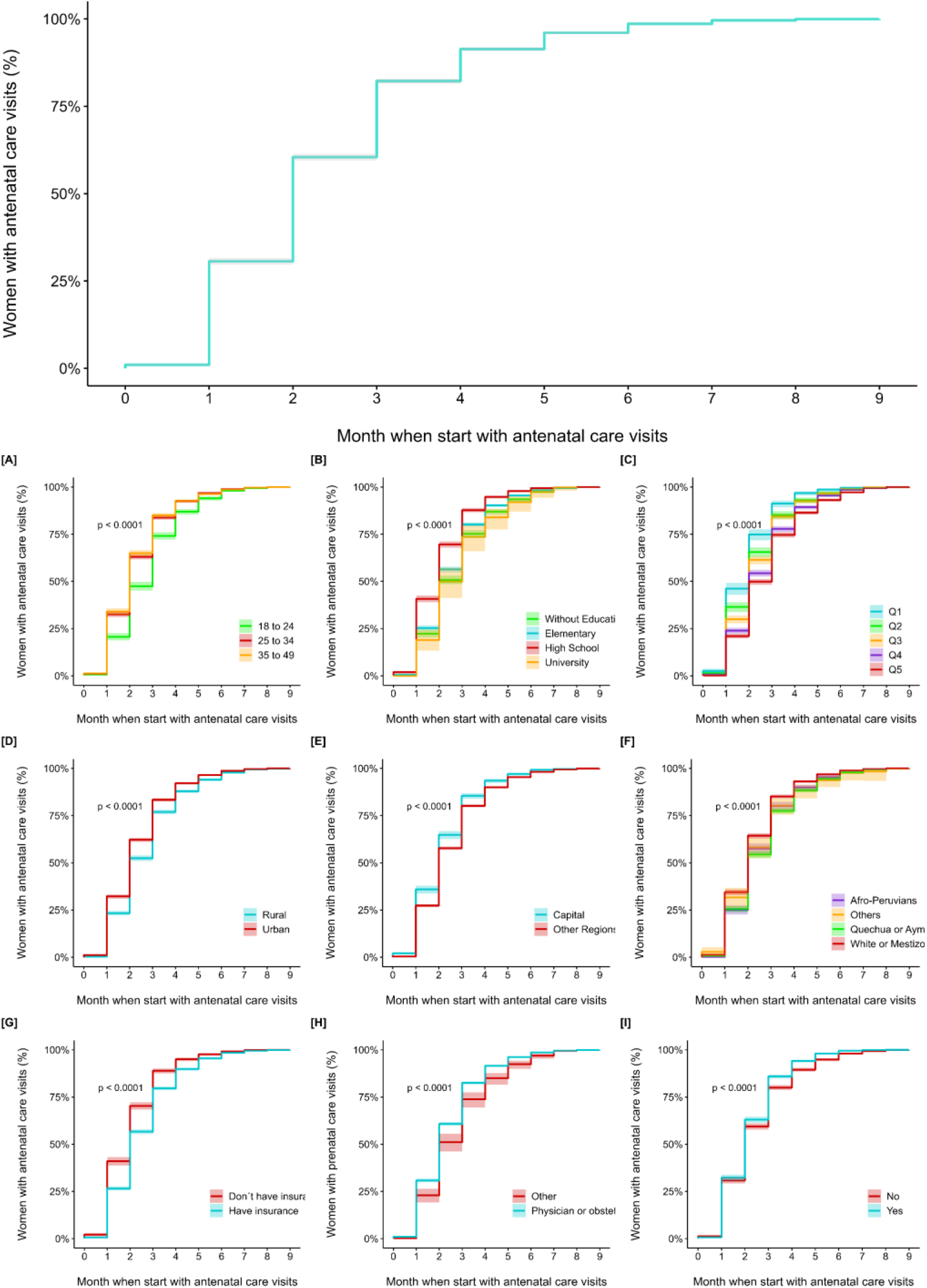
Kaplan-Meier plots to estimate the time elapsed until the beginning of antenatal care visits to the characteristics of Peruvian women of childbearing age

In the survival analysis, the lower educational level and wealth quintile, had lower probability of starting antenatal care visits in the first months of pregnancy, compared to those with higher education and belonging to the first wealth index (Table 2). Likewise, those women living in urban areas had a 6.50% probability of not starting their antenatal care visits in the first months of pregnancy (HRa: 0.94, 95%CI: 0.89 to 0.98, p=0.003), compared to those living in rural areas. In addition, those women who identified as Quechua or Aymara had a 9.10% probability of not starting their antenatal care visits in the first months of pregnancy (HRa: 0.91, 95%CI: 0.87 to 0.95, p<0.001), compared to those who identified as White or Mestizo. Similarly, those women enrolled in comprehensive health insurance during their gestation had a 7.40% probability of not starting their antenatal care visits in the first months of pregnancy (HRa: 0.93, 95%CI: 0.88 to 0.98, p=0.004). Finally, those who received a complete evaluation at their antenatal care visits had a 13.31% probability of starting their antenatal care visits in early pregnancy (HRa: 1.13, 95%CI: 1.09 to 1.18, p<0.001).

**Table 2.** Evaluation of sociodemographic and health characteristics associated with the month of beginning of antenatal care visits in Peruvian women.

| Characteristics | Crude Model |  |  | Adjusted Model |  |  |
| --- | --- | --- | --- | --- | --- | --- |
|  | cHR | 95%CI | P-value* | aHR | 95%CI | P-value* |
| **Age Group?** |  |  |  |  |  |  |
| 18 to 24 years old |  | **REF** |  |  | **REF** |  |
| 25 to 34 years old | 1.26 | 1.21 to 1.30 | <0.001 | 1.22 | 1.16 to 1.28 | <0.001 |
| 35 to 49 years old | 1.28 | 1.23 to 1.33 | <0.001 | 1.26 | 1.19 to 1.33 | <0.001 |
| **Educational Level?** |  |  |  |  |  |  |
| University |  | **REF** |  |  | **REF** |  |
| High School | 0.79 | 0.76 to 0.82 | <0.001 | 0.85 | 0.81 to 0.89 | <0.001 |
| Elementary | 0.72 | 0.69 to 0.75 | <0.001 | 0.79 | 0.74 to 0.84 | <0.001 |
| Without education | 0.69 | 0.62 to 0.77 | <0.001 | 0.74 | 0.63 to 0.85 | <0.001 |
| **Wealth Index?** |  |  |  |  |  |  |
| First Quintile (Q1) |  | **REF** |  |  | **REF** |  |
| Q2 | 0.82 | 0.77 to 0.88 | <0.001 | 0.91 | 0.84 to 0.99 | 0.028 |
| Q3 | 0.78 | 0.73 to 0.83 | <0.001 | 0.89 | 0.82 to 0.96 | 0.003 |
| Q4 | 0.68 | 0.65 to 0.73 | <0.001 | 0.84 | 0.77 to 0.91 | <0.001 |
| Last Quintile (Q5) | 0.63 | 0.6 to 0.67 | <0.001 | 0.81 | 0.74 to 0.88 | <0.001 |
| **Residence Area?** |  |  |  |  |  |  |
| Urban |  | **REF** |  |  | **REF** |  |
| Rural | 1.17 | 1.14 to 1.21 | <0.001 | 0.94 | 0.89 to 0.98 | 0.003 |
| **Do you live in the Capital?** |  |  |  |  |  |  |
| No |  | **REF** |  |  | **REF** |  |
| Yes | 1.14 | 1.1 to 1.19 | <0.001 | 1.04 | 0.99 to 1.08 | 0.129 |
| **Ethnic Group?** |  |  |  |  |  |  |
| White or Mestizo |  | **REF** |  |  | **REF** |  |
| Quechua or Aymara | 0.84 | 0.81 to 0.87 | <0.001 | 0.91 | 0.87 to 0.95 | <0.001 |
| Afro-Peruvians | 0.88 | 0.84 to 0.92 | <0.001 | 0.98 | 0.93 to 1.04 | 0.520 |
| Others | 0.86 | 0.77 to 0.96 | 0.006 | 1.03 | 0.93 to 1.14 | 0.608 |
| **Were you affiliated to SIS during pregnancy?** |  |  |  |  |  |  |
| No |  | **REF** |  |  | **REF** |  |
| Yes | 0.78 | 0.75 to 0.82 | <0.001 | 0.93 | 0.88 to 0.98 | 0.004 |
| **Was the ACVs performed by a physician or obstetrician?** |  |  |  |  |  |  |
| No |  | **REF** |  |  | **REF** |  |
| Yes | 1.20 | 1.12 to 1.28 | <0.001 | 1.02 | 0.94 to 1.11 | 0.598 |
| **Did you have ACVs with complete evaluations?** |  |  |  |  |  |  |
| No |  | **REF** |  |  | **REF** |  |
| Yes | 1.13 | 1.09 to 1.18 | <0.001 | 1.13 | 1.09 to 1.18 | <0.001 |

**cHR:** crude Hazard Ratio, **HRa:** adjusted Hazard Ratio, **95%CI:** 95% Confidence Interval, **ACVs:** Antenatal Care Visits.
\*Estimated p-value using a weighted Cox regression model for a complex sample.

In the analysis of inequality in the month of starting antenatal care visits, it was found that among women who started in the first month of pregnancy, some characteristics such as being 35 to 49 years old (SII: 0.31, 95%IC: 0.24 to 0.37, p<0.001), having a university education (SII: 0.83, 95%IC: 0.80 to 0.86, p<0.001), living in urban areas (SII: 0.75, 95%CI: 0.71 to 0.78, p<0.001) or in the capital region (SII: 0.65, 95%CI: 0.60 to 0.70, p<0.001), identifying as white or mixed race (SII: 0.42, 95%CI: 0.37 to 0.48, p<0.001), and not being affiliated with comprehensive health insurance during pregnancy (SII: 0.77, 95%CI: 0.73 to 0.81, p<0.001), mediated less inequality. Conversely, factors such as age between 18 to 29 years (SII: -0.22, 95%CI: -0.26 to -0.18, p<0.001), 25 to 34 years (SII: -0.09, 95%CI: -0.16 to -0.02, p=0.009), no education (SII: -0.03, 95%CI: -0. 04 to -0.02, p<0.001) or elementary (SII: -0.42, 95%CI: -0.46 to -0.38, p<0.001) and secondary education (SII: -0.53, 95%CI: -0.58 to -0.48, p<0.001), living in rural areas (SII: -0.75, 95%CI: -0.78 to -0.71, p<0.001) or outside of the capital (SII: -0.65, 95%CI: -0.70 to -0.60, p<0.001), identifying as Quechua or Aymara (SII: -0.25, 95%CI: -0.30 to -0.19, p<0.001), Afro-Peruvian (SII: -0.15, 95%CI: -0.18 to -0.11, p<0.001) and being affiliated with national health insurance or Integral Health Insurance (SIS, acronym in Spanish) during pregnancy (SII: -0.77, 95%CI: -0.81 to -0.73, p<0.001) mediated greater inequality for initiating antenatal care visits in the first month of pregnancy.

The pattern of inequality persisted in begining of antenatal care visits in the second, third, and fourth months of pregnancy (Figure 3). Similarly, inequalities were found for initiating antenatal care visits in the first to ninth month of pregnancy if a complete evaluation was performed at these visits. Inequalities also existed in starting antenatal care visits at any point during pregnancy based on the health professional who attended (Figure 3).

**Figure 3.**
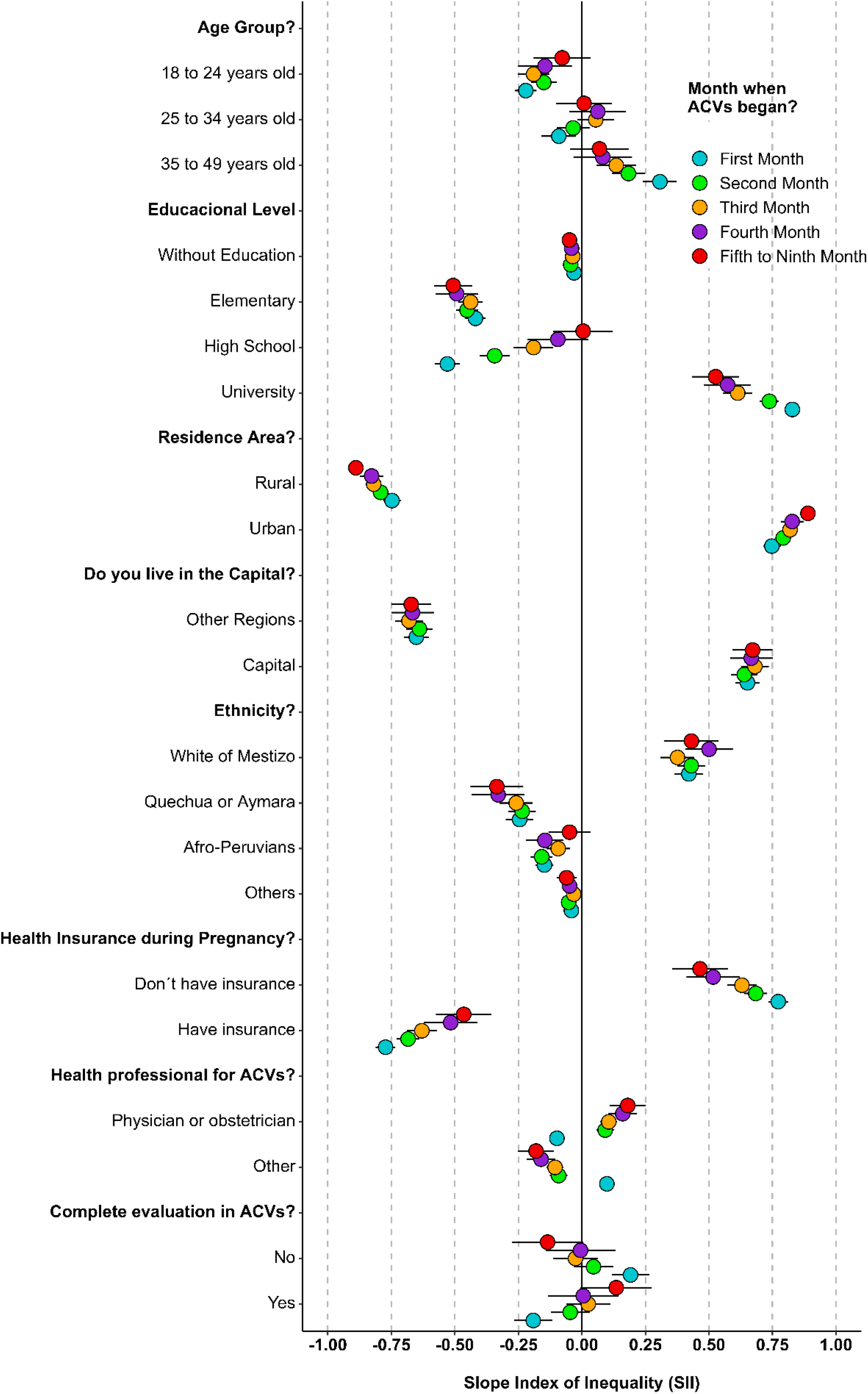
Inequalities related to the month of pregnancy when starting prenatal checkups according to the sociodemographic characteristics of Peruvian women of childbearing age **ACVs:** Antenatal Care Visits.

## DISCUSSION

This study assess sociodemographic inequalities in the early beginning of antenatal care visits during pregnancy among Peruvian women. Most of them began the antenatal care visits around the third month of pregnancy, which varied depending on sociodemographic factors. Therefore, it was found that pregnant women living in rural areas or outside of the capital faced greater inequalities in initiating antenatal care visits during the first five months of pregnancy (2). The study reveals geographic inequalities in Peruvian antenatal care, showing that 25.8% of women in rural areas did not start their antenatal care visits in the first trimester (3). This is due to Peru’s centralization of resources and health services, which limits the availability of physicians or obstetricians in rural areas (4). However, care from these professionals helps to improve the early antenatal care initiation (17).

The contradiction of finding that women affiliated to SIS during pregnancy faced greater inequality for the onset of antenatal care visits in the first five months, this represents how the greater coverage of affiliates does not guarantee early antenatal care visits (18,19). Health care providers in some regions face multiple challenges in delivering early care to pregnant women, particularly in rural areas and outside of Lima (2,4). And even when in Peru, antenatal care visits are performed in primary health care centers, the lack of focus on the inequalities in pregnant women affiliated with the national health insurance may result in a delayed onset of antenatal care visits, leading to increased risks of developing comorbidities and complications in labor and delivery (20,21).

Pregnant women identified as Quechua or Aymara, Afro-Peruvian, or other non-white or mestizo ethnic groups showed greater inequality in the onset of antenatal care visits. This is a consequence of adverse socioeconomic conditions among indigenous ethnic groups in Peru, which decrease access to health care services (22). In addition, language gaps and experiences of discrimination mitigate the intention to seek antenatal care, even more so when these groups are more likely not to have quality antenatal care visits (23). Therefore, intercultural health strategies for pregnant women could improve early initiation, compliance with the minimum number of visits, and the quality of antenatal care for various ethnic groups (2,3).

Also, it was found that pregnant women over 35 years of age were more likely to have an early onset of antenatal care visits. This is because older pregnant women are at greater risk of developing comorbidities, so the concern for a healthy pregnancy may condition the onset of antenatal care visits in the first months (24). On the other hand, it was found that pregnant women between 18 and 24 years of age had a greater inequality in the initiation of antenatal care visits in the first four months of pregnancy compared to older pregnant women. Among aged women is more probably a stable job and family support, which allows them to plan pregnancy and access health care services compared to younger women (7,25). This finding is reinforced by the lesser inequality faced by college-educated women, who are more likely to understand the importance of pregnancy care and potentially have better access to a greater variety and quality of antenatal care services (26).

Pregnant women who received the evaluations established as standard of care in antenatal care visits were more likely to have an early onset in the sessions during the first months of pregnancy. As a result, less than half of the pregnant women who initiated their antenatal care visits in the first month received complete antenatal care evaluations (15). This underscores the necessity of educating pregnant women about the risks of contracting illnesses before or during pregnancy, as well as the critical importance of receiving supplementation for optimal fetal development (27). In addition, providing information on healthy eating, complications, and emergency care during pregnancy allows for greater involvement of pregnant women in achieving a better labor and delivery (28).

This research provides an approximation of inequalities in the early initiation of antenatal care visits. However, limitations stemmed from potential biases in data collection by the DHS in Peru, where some women may have omitted details about their pregnancy (recall bias) or provided socially desirable responses, affecting the study’s accuracy. The lack of follow-up on pregnant women in Peru through national surveys impedes a comprehensive understanding of how these inequalities influence pregnancy outcomes and the health of newborns. On the other hand, the period covered by the study includes the years in which COVID-19 impacted Peru, which could have a considerable impact on the estimates made; however, since there is no information on the year of pregnancy or whether pregnant women was infected with SARS-CoV-2, it is difficult to estimate the influence of the pandemic in the study.

In conclusion, this study sheds light on the significant sociodemographic inequalities at the beginning of antenatal care visits among Peruvian women, particularly highlighting the challenges faced by those residing in rural areas or outside the capital, without a university education, and those identified as Quechua or Aymara. The findings underscore the geographical inequalities entrenched within Peru’s healthcare system, where limited access to healthcare professionals in rural regions contributes to delayed initiation of antenatal care, exacerbating risks during pregnancy and childbirth.

## Data Availability

All data produced are available online at: https://proyectos.inei.gob.pe/microdatos/

## Conflict of interests

None.

## Funding

Self-funded

## Acknowledgements

None.

### APPENDICES

**Appendix 1.**
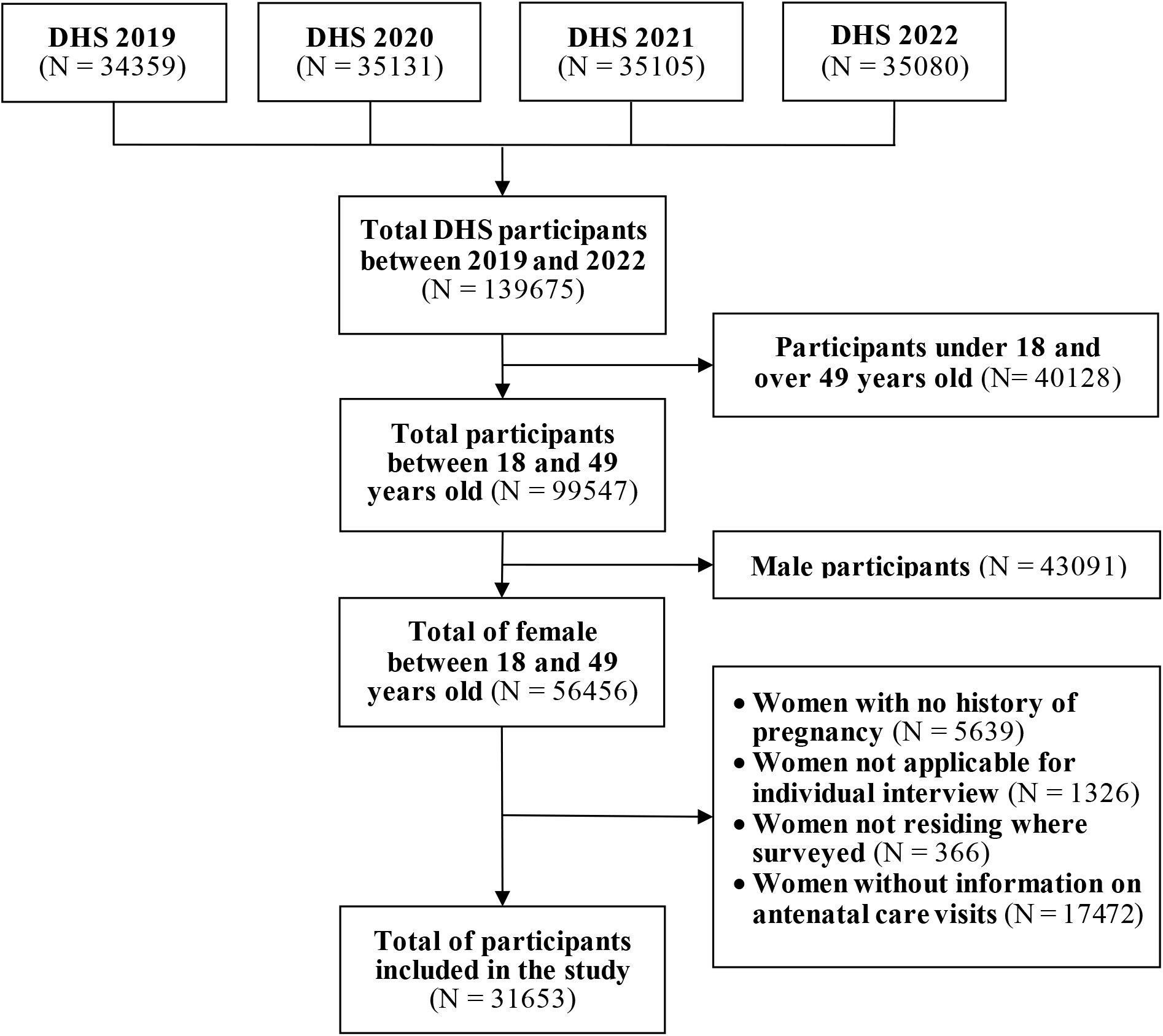
Flowchart of selection of participant in DHS between 2019-2022 **DHS:** Demographic and Family Health Survey in Peru.

## Notes

### Competing Interest Statement

The authors have declared no competing interest.

### Funding Statement

This study did not receive any funding

### Author Declarations

A request for ethical committee evaluation was not necessary as the data collection process for Demographic Health Survey in Peru was carried out with the informed consent of participants. Furthermore, the data obtained from the National Institute of Statistics and Informatics (or INEI, in Spanish) platform does not contain personally identifiable information and is secure to be used, this data is located at: https://proyectos.inei.gob.pe/microdatos/

